# Plasma proteins identify increased number of carotid plaques and predict occurring atherosclerotic events

**DOI:** 10.1101/2023.09.11.23295388

**Authors:** A Baragetti, L Grigore, E Olmastroni, E Mattavelli, AL Catapano

## Abstract

**Background:** The number of carotid plaques independently predicts incident atherosclerotic cardiovascular disease (ACVD).

However, performing vascular imaging in apparently healthy subjects is challenging, owing organizational/economical barriers. Plasma proteomics can offer an alternative approach to identify individuals with carotid plaques, at high risk of eventually developing ACVD.

**Methods:** In this observational study, we studied by Normalized Protein eXpression (NPX; Olink^TM^), the plasma levels of 368 proteins in 664 subjects from the PLIC study, who were screened by ultrasound for the presence of carotid plaques. We clustered, by artificial intelligence, the proteins that more accurately identified subjects, stratifying them according to the number of plaques they presented with. We also study prediction of occurring events over 22 years.

**Results:** 299/664 subjects had at least 1 carotid plaque. Among those, 77 subjects presented with only one plaque, 101 with 2 plaques and 121 with ≥3 plaques (3+). The remaining 365 subjects with no plaques acted as controls. The proteins differently expressed versus controls increased as a function of the number of plaques. 32 proteins were shared among the groups of subjects with plaques, but 87, significantly associated with the presence of 3+ plaques, improved the AUC of the ROC, together with the ACVD risk factors, to discriminate subjects with 3+ plaques versus the AUC of the ROC considering the ACVD risk factors only (AUC= 0.918 (0.887-0.943) vs AUC= 0.760 (0.716-0.801) respectively, p<0.001). The ACVD risk factors barely predicted the 198 occurring events (AUC= 0.559 (0.521-0.598)), but proteomics associated with plaques improved the prediction (AUC= 0.739 (0.704-0.773), p<0.001).

By analyzing the biological processes, we identified that chemotaxis/migration of leukocytes and the signaling of interleukins/cytokines were the top pathways involved.

**Conclusions:** Plasma proteomics helps to identify apparently healthy subjects with higher number of carotid plaques more accurately and to predict occurring ACVDs in those individuals.

## Introduction

Atherosclerotic Cardiovascular Disease (ACVD) is the leading cause of death worldwide^1^. In “apparently healthy” population a subset of subjects with subclinical ACVD can be detected and these subjects are identified as high, or very high, risk in contemporary cardiovascular guidelines, and carotid plaques have been suggested as a hallmark of increased ACVD risk ^2–4^.

However, applying noninvasive vascular imaging procedures to detect subclinical ACVD, such as those identifying carotid plaques, is challenging in “apparently healthy” population, owing organizational, economical barriers, and given the substantial shortcomings in sensitivity and inter-operator variability ^5,6^.

Plasma biomarkers have been recently used to identify people at high risk of developing^7^ or reoccurring cardiovascular events^8^ and accumulating evidence indicates that plasma proteins, or specific subsets of them, outperform the currently available algorithms to predict increased ACVD risk in “apparently healthy” individuals^7^. Yet evidence regarding their ability to inform the presence of vascular plaques is still limited, as in few studies the number of plasma proteins that have been investigated is limited to 86 ^9,10^.

Inflammation is known to play an important role in the onset and progression of plaques^11^. We therefore sought to address the question whether we could more accurately identify “apparently healthy individuals” carrying asymptomatic carotid plaques among 664 subjects from a free-living population of the northern area of Milan. To this end we studied, by Normalized Protein eXpression (NPX) (an index of relative abundance^12^), the plasma levels of 368 proteins from the targeted Cardio-metabolic, Inflammation, Cardiovascular II, and III panels (Olink^TM^). New omics techniques can inform on vascular activation of pathway related to atherosclerosis^13–15^ and the power of proteomics as an independent marker of carotid atherosclerosis has been still proposed, albeit in a limited number of subjects and by addressing a maximum of 82 plasma proteins linked to cardiovascular disease ^16,17^. Therefore, we searched for proteins whose expressions in plasma, recapitulating biologically relevant pathways, can improve the accuracy to identify apparently healthy subjects with asymptomatic atherosclerotic disease at the carotids and if they also could predict the occurrence of clinically overt atherosclerotic stages.

## Materials and methods

### Study population

The population was from a subgroup of subjects from the northern area of Milan, randomly selected during the basal clinical evaluation of the Progressione delle Lesioni Intimali Carotidee (“PLIC”) Study, which initially included 2.606 participants from 2001 to 2003 ^18,19^. Targeted plasma proteomics was available for 702 participants, but only 664 subjects, who did not develop ACVD before the collection of plasma, were considered for the purpose of the study (**Supplemental Figure 1**). Out of them, 365 were free of carotid plaques detected by ultrasound and acted as controls. The subjects’ characteristics agreed with the Sex and Gender Equity in Research (SAGER) guidelines ^20^. The PLIC Study has been approved by the Ethics Committee of the University of Milan (SEFAP/Pr.0003). Written informed consent was obtained from subjects (all over 18 years-old), in accordance with the Declaration of Helsinki. This work is an observational study and it was conducted following the standards of the STrengthening the Reporting of OBservational studies in Epidemiology (STROBE) initiative^21^.

### Clinical and biochemical data

Participants in this study were screened at the Center for the Study of Atherosclerosis at E. Bassini Hospital (Cinisello Balsamo, Milan, Italy) for the personal, familial, pathological, and pharmacological clinical history. Subjects with malignancies (any type), chronic liver disease (self-reported or clinically documented in the records of the Hospital), chronic kidney disease (Glomerular Filtration Rate, GFR < 60 ml/min or documented albuminuria > 30 mg/g) and pregnant women were excluded from the study.

Information about lifestyle habits (physical activity, former or active smoking habit) was also collected by validated questionnaire used in the PLIC Study as described elsewhere^22^. Systolic and diastolic blood pressure, body mass index (BMI), waist and hip circumferences were measured. We followed the recent EAS/ESC guidelines for the diagnosis of hypertension, for the diagnosis of dyslipidemia and for the definition of the cut-off of waist circumference (>94 cm for men and >88 cm for women)^23^. Blood samples were collected from antecubital vein after 12 hours fasting on NaEDTA tubes (BD Vacuette), centrifuged at 3,000 rpm for 12 minutes (Eppendorf 580r, Eppendorf, Hamburg, Germany) aliquoted in multiple 200 ul samples, stored at -80 degrees for the subsequent profiling of biochemical parameters (which included: total cholesterol, High-Density Lipoprotein cholesterol (HDL-C), triglycerides, ApoB, ApoA-I, glucose, Lp(a), and C-Reactive Protein (CRP)). All the measurements were performed on the aliquots, thawed for the first time, using immuno-turbidimetric and enzymatic methods thorough automatic analyzers (Randox, Crumlin, UK). Low-Density Lipoprotein cholesterol (LDL-C) was derived from Friedewald formula. Lipoprotein(a) (Lp(a)) was measured by immunoassay (Randox, Crumlin, UK).

### Carotid ultrasound

Carotid ultrasound analysis was performed by a single expert sonographer, blinded on subject’s identity and clinical history, with a linear ultrasound probe (4.0e13.0 MHz frequency, 14X48 mm footprint, 38 mm field of view, Vivid S5 GE Healthcare®, Wauwatosa, WI, USA). A detailed description of the ultrasound methodology, based on validated protocol for the PLIC study, has been published elsewhere^19^.

Carotid plaques were defined if (i) focal loci were identified and/or (ii) if the measured carotid Intima Media Thickness (IMT) was equal or above than1.3 mm in any vascular tract (common, bulb, bifurcation, internal and external tracts) of one or both carotid arteries ^24^.

### Plasma proteomics

368 proteins were measured in plasma by Proximity Extension Assay (PEA) technology. The proteins are included in the Cardiovascular II, Cardiovascular III, Inflammation and Cardio-metabolic panels of Olink^TM^ platform. The complete list of the proteins that are included in the panels, their Uniprot ID and the Olink ID, are provided by the company (http://www.olink.com). Proteins were measured from 200 uL of plasma samples by Proximity Extension Assay (PEA). Plasma samples from each subject was separated from whole blood by centrifugation (3,000 rpm, 12 minutes at 4 degrees), aliquoted in 96-wells plates during the first clinical evaluation of each subject (between 1998 and 2000) and immediately stored at -80 degrees up to 2018, when they were sent to Olink^TM^ (Uppsala, Sweden) for proteomic characterization ^7^. The final assay read-out of each determination is given in Normalized Protein eXpression (NPX), which is an arbitrary unit on log2-scale where a high value corresponds to a higher protein plasma level. Each PEA measurement has a lower detection limit (LOD) calculated versus negative controls that are included in each run, and measurements of individual proteins below LOD for each panel were removed from further analysis (2 samples from Cardiometabolic panel, 29 samples from Cardiovascular II; 31 samples from the Inflammation panel; all the samples were with measurement>LOD in the Cardiovascular III panel). All assay characteristics including detection limits, the measurements of assay performance, and validations are available from the manufacturer webpage (http://www.olink.com). All raw data can be provided upon reasonable written request to ALC.

### Statistical analysis

#### Descriptive statistics

Clinical and biochemical data are presented as median with interquartile ranges (the lower 25th percentile and the upper 75th percentile). After checking for the type of distribution of linear variables (Shapiro-Wilk Test), we compared them between either two groups (Kolmogorov-Smirnov test) or four groups (controls, subjects with 3+, with 2 and with 1 carotid plaque), either by Analysis of Variances (ANOVA) and Bonferroni post-hoc analysis (if variables were normally distributed variables), or by Kruskal-Wallis test (if non-normally distributed variables). Dichotomous variables were compared by Fisher’s chi-square test. Grubb’s test for the detection of the outliers was performed for all the linear variables before running tests. P values less than 0.05 were considered significant for all the statistical analyses. SPSS^TM^ v.24 for Windows (IBM corporation®) was used for all statistical analyses. Graphs were prepared using Graphpad® (v.9.0) and Excel (Microsoft®).

#### Machine Learning model

Proteomics data were analyzed by Machine Learning (ML) model, previously published for proteomics data in PLIC cohort^25^.

In brief, the ML model consists of an eight binary outcome gradient boosting prediction model (XGboost), including the 368 proteins measured in the cohort. The XGboost model is a statistical learning technique, which produces a non-linear model in the form of an ensemble of weak prediction tree-based models. The extreme gradient boosting classification algorithm optimizes a cost function^26^ by iteratively choosing a weak hypothesis that points in the negative gradient direction. Using a fivefold cross-validation by random reshuffling of the training set, overfitting was avoided. The XGBoost classifier model was trained in the derivation cohort with 1000 iteration round and 0.300 learning rate. Hyperparameter optimization was performed by k-fold iteration. To avoid overfitting of the models, the dataset was randomly split into a training set (70% of total sample; “train set”) and the algorithm was thereafter applied to the internal validation set (30% of total sample; “test set”). We applied a permutation (randomization test) to evaluate statistical validity of the results, since standard univariate significance tests cannot be applied to the used models due to the large number of tests. By evaluating the distribution of all the results obtained in these simulations and comparing it to the true outcomes, we computed statistical significance associated with the joint panel of the selected markers. We also reported importance scores for each of the proteins that demonstrate preferences of model when constructing non-linear prediction function based on the selected biomarkers. (Random Forest classifiers). Sensitivity and specificity to predict the outcome, were performed by running analysis of Receiver Operating Curves (ROC). Comparison of AUC was performed according to the method of Hanley-McNeil. All the ML models were constructed by using Python 6.4.5 with pandas, scikit–learn, NumPy, XGboost.

#### Gene Ontology (GO) and KEGG pathway enrichment analyses

We conducted enrichment analysis of biological processes with the proteins that emerged as significantly associated with carotid atherosclerosis following in silico strategies, as previously published ^27,28^. The DAVID (The Database for Annotation, Visualization, and Integrated Discovery, NIAID, North Bethesda, MD, USA) platform was used for gene ontology (GO) enrichment analyses. The significant GO biological processes (GO_bp) were selected for Pvalue<0.05. Then, for each GO biological process (GO_bp) we annotated the fold of enrichment, an index of the percentage of proteins belonging to a pathway, and the false discovery rate (FDR) to indicate how likely the enrichment is by chance (FDR<0.05 indicates a statistically significant enrichment of proteins in that pathway).

## Results

### Increased number of plasma proteins associate with a higher number of carotid plaques

Out of 664 apparently healthy subjects, 299 presented with at least one plaque in one of the carotids, while 365 did not present any plaque and acted as “controls” (the clinical and biochemical parameters are reported in **Supplemental Table 1**). Out of the 299 subjects presenting at least one plaque, 77 had one, 101 subjects had 2 and 121 subjects presented with 3 or more plaques (by 3 up to 7 plaques as a maximum; herein defined as “3+”; the ultrasonographic results are reported in **Supplemental Table 2**). The analysis of the prevalence of the main ACVD risk factors showed a significant trend for active smoking, hypertension, dyslipidemia, increased waist circumference and reduced physically activity associated with an increased number of plaques (**Supplemental Table 3**). Of note, the plasma levels of a widely used marker of chronic low-grade inflammation, CRP, did not differ among all the groups (P_trend_=0.427 **Supplemental Table 3**), while the number of plasma proteins with significantly different NPX increased as a function of the number of plaques vs controls. In fact, 17 proteins were significantly different in subjects with 1 carotid plaque compared to controls (1 reduced and 16 increased; **Figure 1A**, **Supplemental Table 4A**), 47 proteins were differently expressed in the plasma of the subjects with 2 plaques (4 proteins reduced and 43 increased; **Figure 1B**, **Supplemental Table 4B**) and 97 proteins were differently expressed in subjects with 3+ plaques versus controls (9 reduced and 88 increased; **Figure 1C**, **Supplemental Table 4C**).

**Figure 1.**
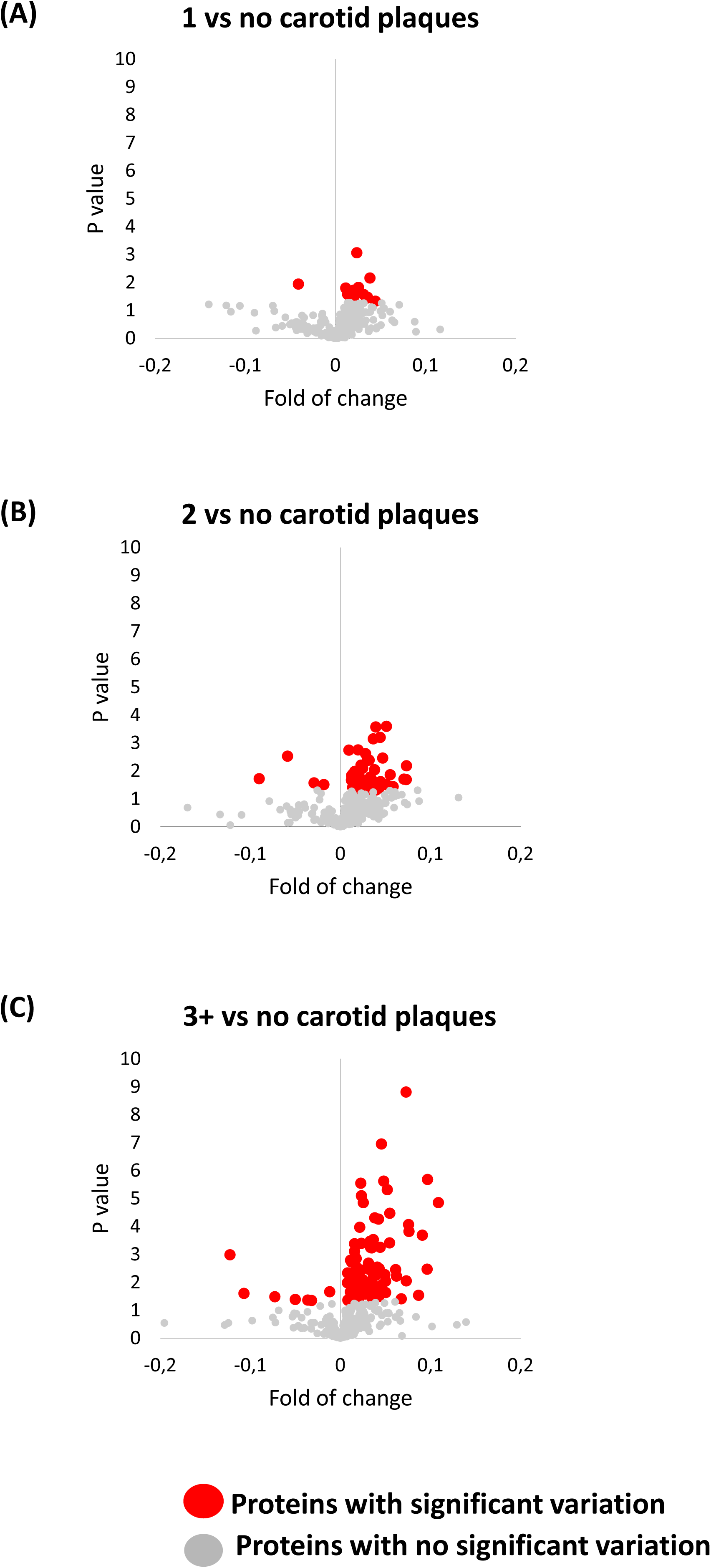
Variations of plasma proteomics between subjects with one, with 2 and with 3+ carotid plaques compared to controls. Volcano plots showing the variation of the 368 plasma proteins in **(A)** subjects with 1 carotid plaque, in **(B)** subjects with 2 carotid plaques and **(C)** in subjects with 3+ plaques. All the variations are referred to controls and are calculated as fold of changes (x axis). In the y axis, the p value is reported indicating the significance of the variation.

Overall, 32 shared proteins were differently expressed among subjects with increasing number of plaques versus controls. Indeed, 6 proteins, increased in subjects with 1 plaque, were also higher in subjects with 2 and in those with 3+ plaques versus controls, the NPX of 2 additional proteins were also higher in subjects with 3+ plaques only (**Table 1**) and, finally, 24 proteins, 2 reduced and 22 increased in subjects with 2 plaques, were present also in subjects with 3+ plaques (**Table 1**).

**Table 1.**
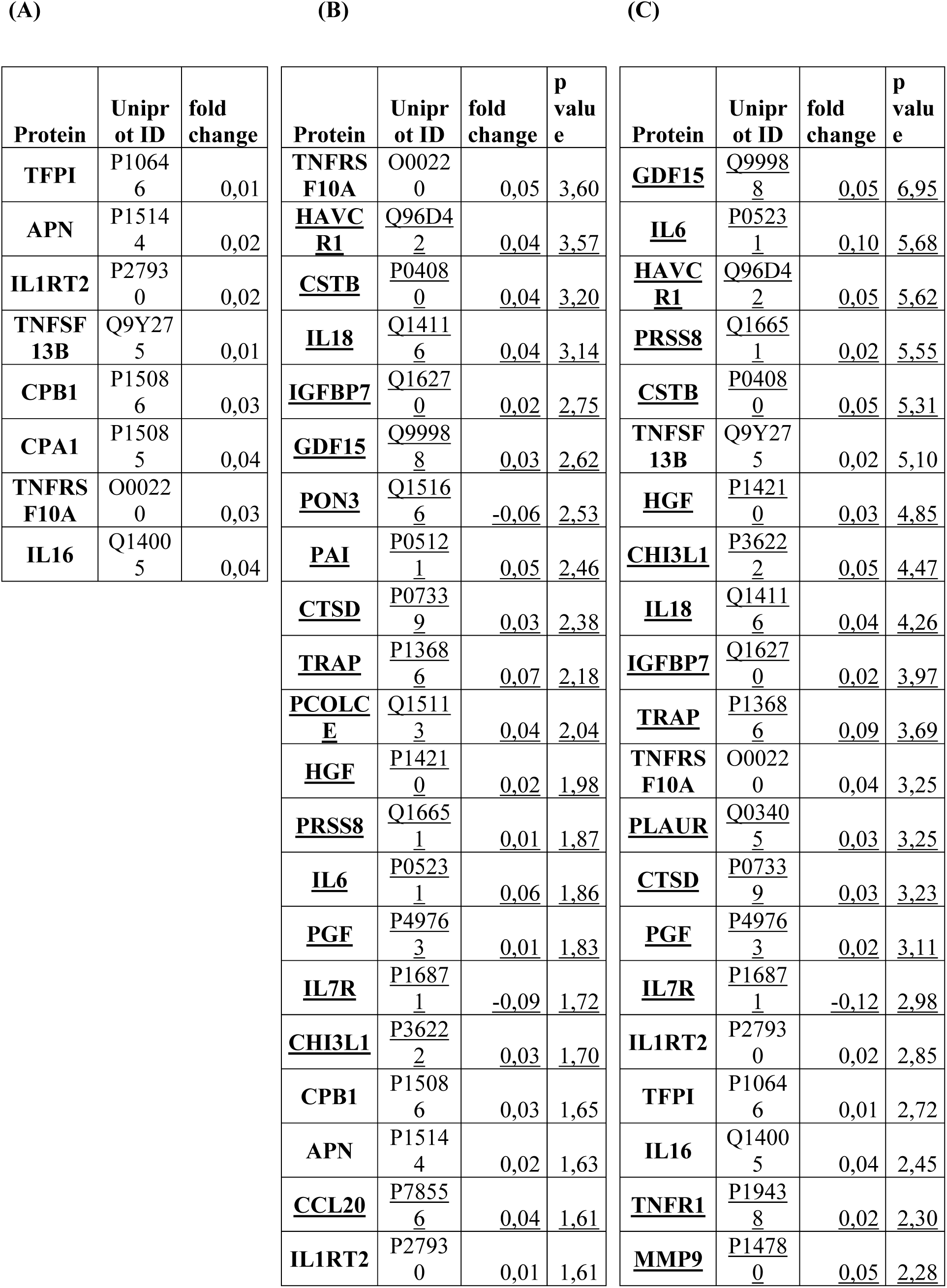
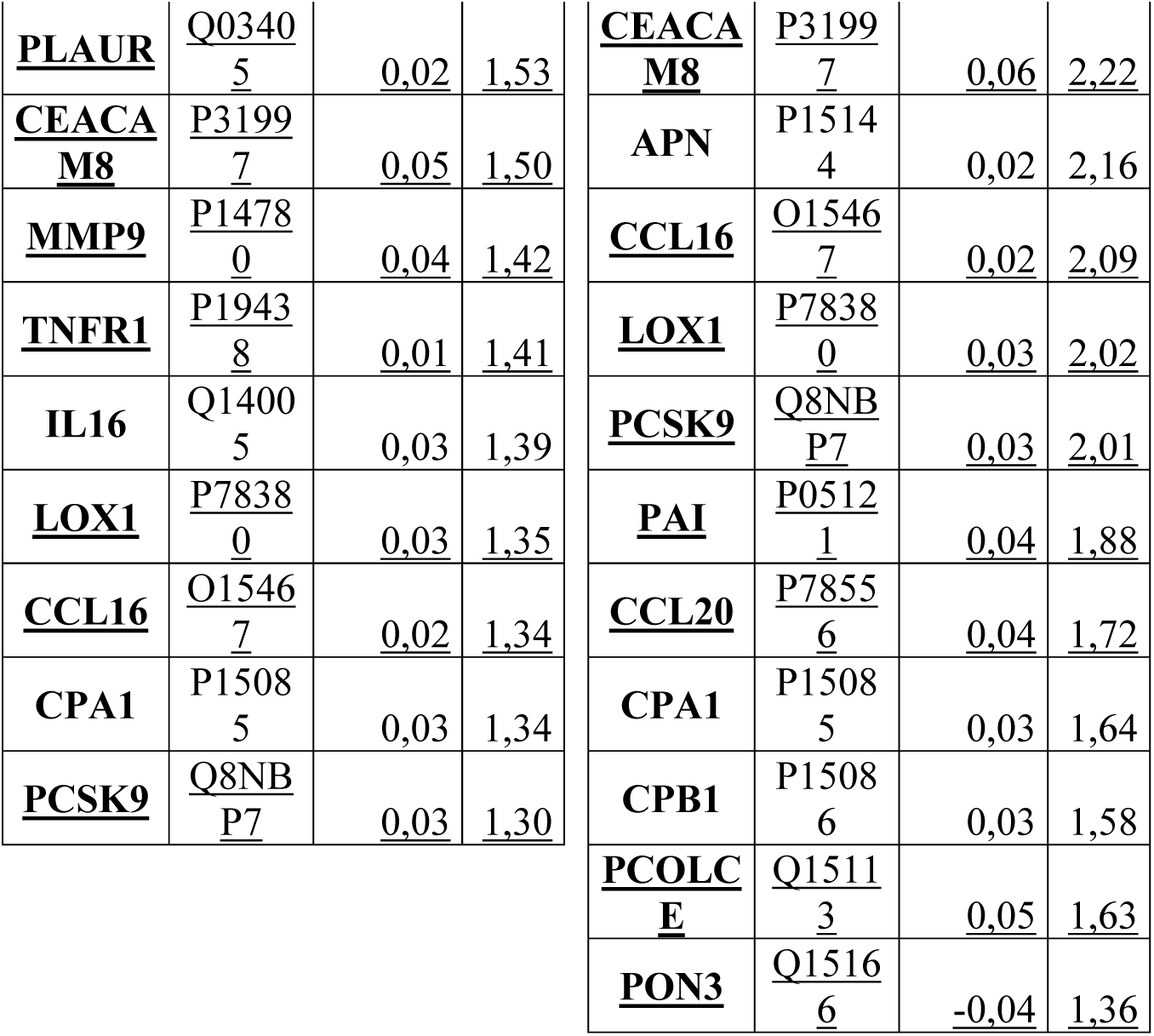
Shared proteins found significantly associated among the subjects with one, two or with 3+ carotid plaques compared to controls. List of proteins with their name, their Uniprot-ID and their fold change in **(A)** subjects with one plaque only, in **(B)** subjects with two plaques and **(C)** in subjects with 3+ plaques. All the fold changes are referred compared to controls. Proteins that are shared between subjects with 2 and subjects with 3+ plaques only are underlined.

### The shared proteins associated with increasing number of plaques recapitulate biological pathways related with atherosclerosis

We analyzed whether the plasmatic expressions of the 32 shared proteins recapitulate biological pathways associated with the development of carotid atherosclerosis. To this end, we performed a protein-protein network (STRING v.11^29^) and gene ontology (GO) enrichment analyses (see methods). By interrogating *in silico* dataset, we predicted that almost all the 32 proteins could be linked into a network of possible biologically relevant pathways, with Interleukin-6 (IL6), Matrix-Metalloproteinase-9, Interleukin-18 (IL18) and plasminogen activator inhibitor 1 (SERPINE1) being central connectors (**Figure 2A**). We then deciphered this complex scenario by gene ontology (GO) enrichment analysis and, although up to 51 representative biological processes were annotated (“GO_bp” in **Supplemental Table 5**), only 9 of them were principally represented by most of the 32 proteins with a significant probability (FDR<0.05; the fold of enrichment for each GO_bp is reported in **Supplemental Table 5**). These GO_bp included pathways biologically relevant for the development of atherosclerosis, such as “proteolysis”, “immune response”, “cytokine-mediated signaling pathway”, “signal transduction”, “cell-cell signaling”, “positive regulation of inflammatory response”, “chemotaxis” and “defense response to Gram positive bacteria” (**Figure 2B**).

**Figure 2.**
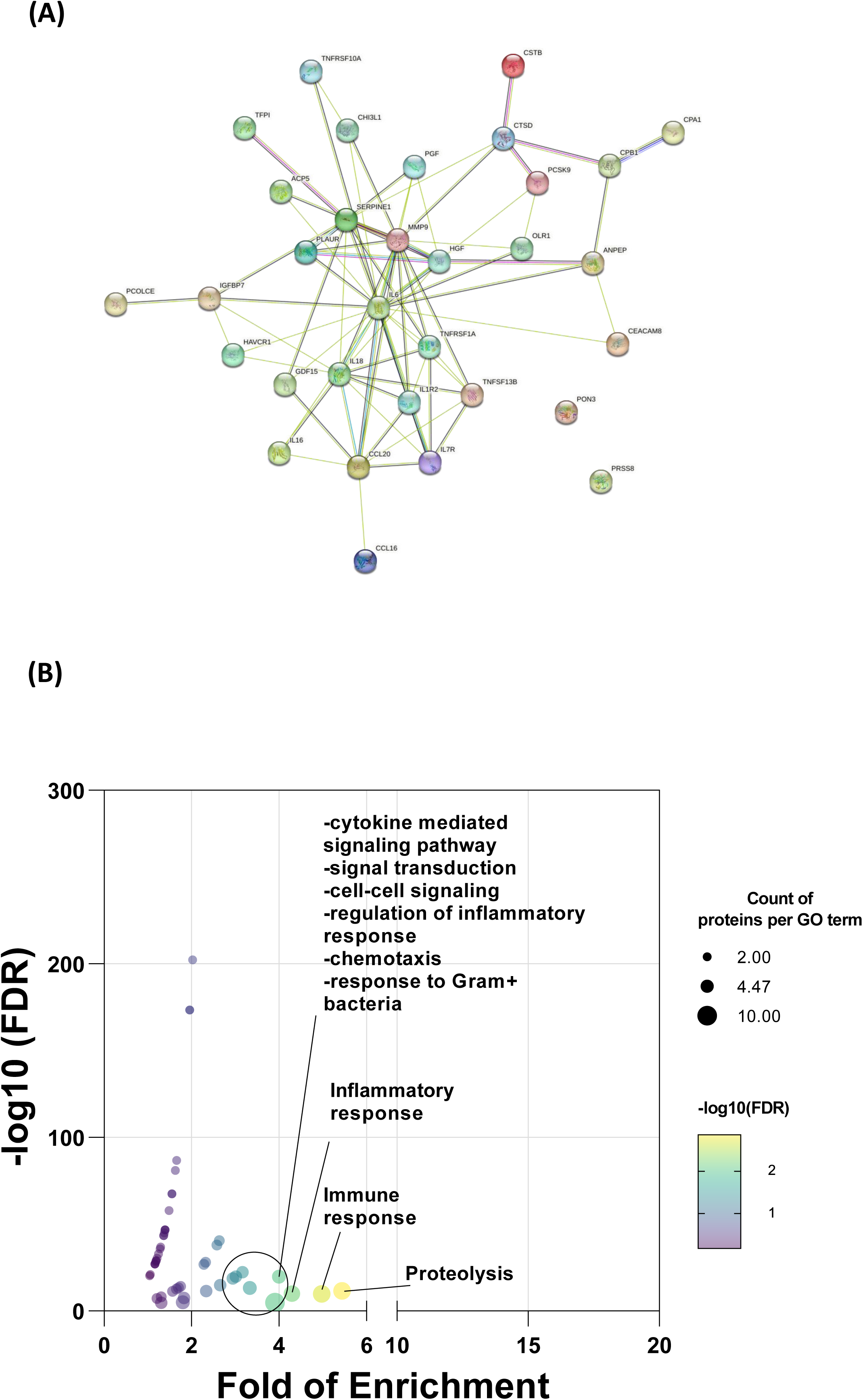
The shared proteins associated with increasing number of plaques recapitulate biological pathways involved in atherosclerosis. **(A)** STRING protein-protein association network reporting the predicted connections between the 32 shared proteins associated with the number of carotid plaques. **(B)** Bubble plot showing the most significant GO_bp processes represented by the 32 shared plasma proteins. Each pathway is plotted for its fold of enrichment and the significance in the intensity (-log 10 FDR) of the signal from proteome analysis (see methods). Significant signals were considered for FDR<0.05.

### A higher number of biological pathways associates with the presence of a higher number of carotid plaques

We also sought to investigate whether the additional proteins that were not shared among the groups of subjects with increasing number of plaques, inform on non-redundant biologically relevant pathways.

Indeed, 9 proteins (out of the 17) were differently expressed only in subjects with 1 plaque; **Supplemental Table 4A**), 17 (out of the 47) only in subjects with 2 plaques; **Supplemental Table 4B**) and 65 (out of the 97) only in subjects with 3+ plaques (**Supplemental Table 4C**).

We found 5 GO_bp that, out of 13 annotated, barely represented the presence of one carotid plaque only compared to controls (-log_10_p-value≥1.3; **Supplemental Table 6A**) and included “viral entry into host cells” and other T-cell mediated pathways (**Figure 3A**). By contrast, the number of GO_bp significantly associated with the presence of 2 plaques only increased up to 22 (out of 35 annotated; **Figure 3B**), with only one of them being also found to be associated with the presence of 1 plaque (“positive regulation of NF-kappaB transcription factor activity”; in **Supplemental Table 6B**).

**Figure 3.**
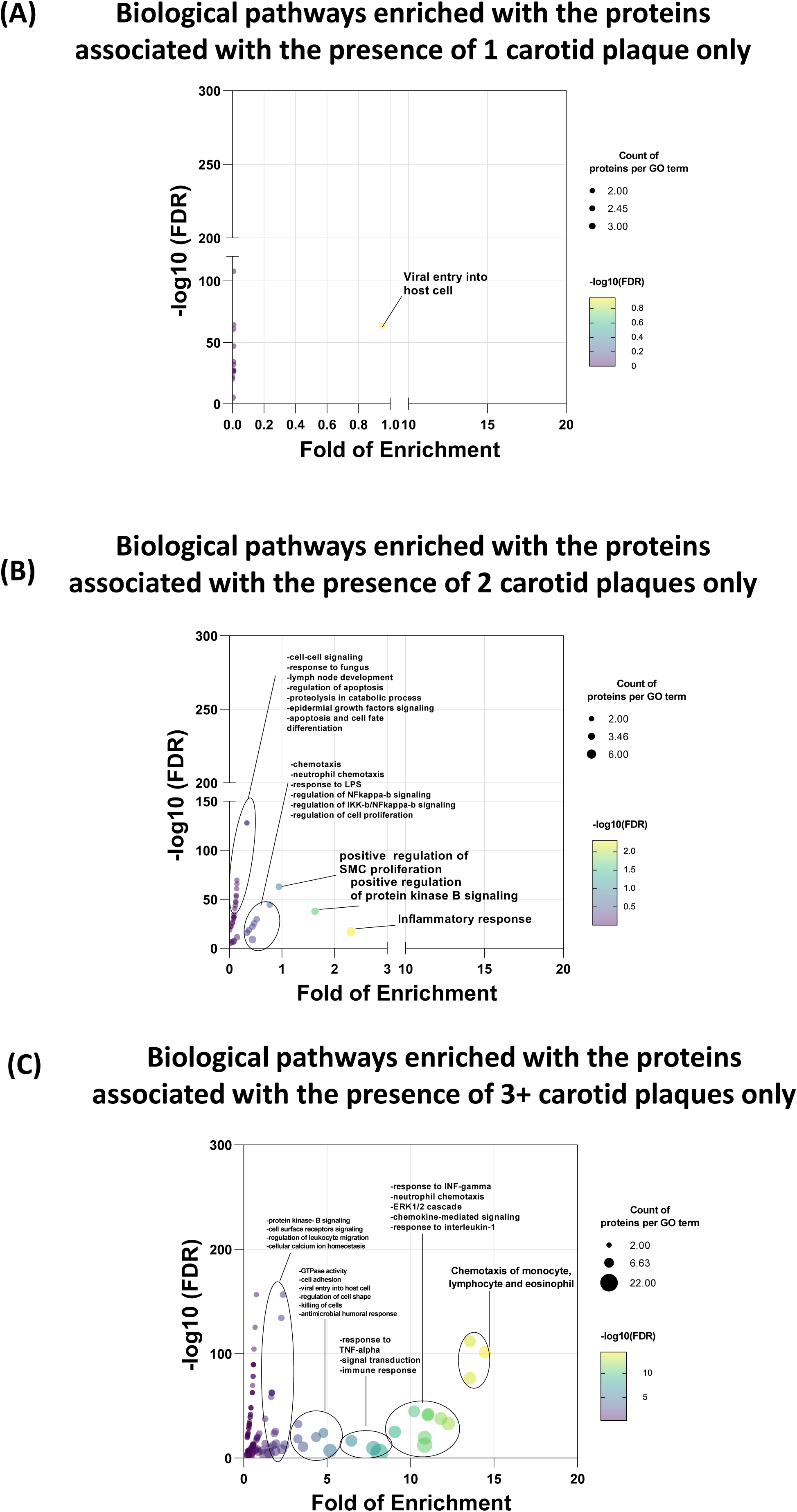
A higher number of biological pathways associates with the presence of a higher number of carotid plaques. Bubble plot showing the most significant GO_bp processes represented by **(A)** the unshared proteins associated the presence of one plaque only, **(B)** the unshared proteins associated the presence of two plaques and **(C)** the unshared proteins associated the presence of 3+ plaques.

Finally, we found that the number of GO_bp significantly associated with the presence of 3+ plaques markedly increased up to 80 (out of 104 annotated; **Figure 3C**). It is of note that only 6 of them were also found among the pathways associated with the presence of one plaque only (“response to hypoxia”, “positive regulation of protein phosphorylation”, “signal transduction”, “T cell co-stimulation”, “positive regulation of peptidyl-serine phosphorylation”, “viral entry into host cell”) and 16 were also found among the pathways associated with the presence of 2 plaques (including pathways involved in leukocytes chemotaxis, migration, cell adhesion, inflammatory response, immune response, and other more specific, such as “positive regulation of MAP kinase activity”, “positive regulation of protein kinase B signaling”, “response to lipopolysaccharide”, “defense response to Gram-bacterium” and “antimicrobial humoral immune response mediated by antimicrobial peptide”) (**Supplemental Table 6C**).

This observation indicates that, especially in the presence of 3+ plaques, an increased number of proteins is linked to a larger number of pathways involved in the development of atherosclerosis.

### Plasma proteins improve the accuracy of predicting the number of carotid plaques

We looked for the performance of plasma proteomics to improve the capability of the classical ACVD risk factors to distinguish the apparently healthy subjects who are still exposed to carotid atherosclerosis.

Only the 97 proteins that were differently expressed in subjects with 3+ plaques versus controls contributed to an improvement of the ROC (AUC= 0.683 (0.601-0.785), p<0.001) (**Table 2**), while neither the 17 proteins that were differently expressed in subjects with only one plaque, nor the 47 differently expressed in subjects with 2 plaques, exerted this performance (AUC=0.504 (0.461-0.591), p=0.948 and AUC= 0.589 (0.503-0.671), p=0.083, respectively (**Table 1**)).

**Table 2.** Targeted plasma proteomics outperforms the identification of carotids with higher number of plaques. Table reporting the parameters of the Receiving Operating Curves (ROC) for the models including the proteins differently expressed either in subjects with one plaque only, or in subjects with 2 or in subjects with 3+ plaques compared to controls. The mean Areas Under the Curve (AUC), the 95% confidence intervals (C.I.), the z-statistics and the p-value for each ROC are reported.

| **Performance of the models to distinguish:** | **AUC** | **Standard error** | **95% C.I. (lower-upper)** | **z-statistic** | **p-value** |
| --- | --- | --- | --- | --- | --- |
| **Number of plaques** |  |  |  |  |  |
| 1 plaque vs controls | 0.504 | 0.0561 | 0.461-0.591 | 0.065 | 0.948 |
| 2 plaques vs controls | 0.589 | 0.0513 | 0.503-671 | 1.735 | 0.083 |
| 3+ plaques vs controls | 0.683 | 0.0471 | 0.601-0.785 | 3.892 | <0.001 |

Next, to find the proteins, out of the 97, whose expression in plasma can significantly improve the accuracy to identify subjects with higher number of carotid plaques on top of the factors commonly considered for the assessment of ACVD risk, we harnessed machine learning model which listed 87 important contributors for a significantly increased performance of the ROC curve. Of note, these 87 proteins improved the accuracy of the classical ACVD risk factors (hypertension, active smoking habit, increased waist circumference, dyslipidemia and reduced physical activity) to identify the subjects with 3+ plaques from controls (**Supplemental Table 7** lists the proteins by descending importance factor derived from random forest classifier and their mean NPX (±standard error), being reduced for 5 and increased for 82 proteins in subjects with 3+ plaques vs controls). Indeed, the AUC of the model including these 87 proteins together with the risk factors was markedly superior to the AUC of the model including the same ACVD risk factors alone (AUC= 0.918 (0.887-0.943) vs AUC= 0.760 (0.716-0.801) respectively, p<0.001; **Figure 4A**).

**Figure 4.**
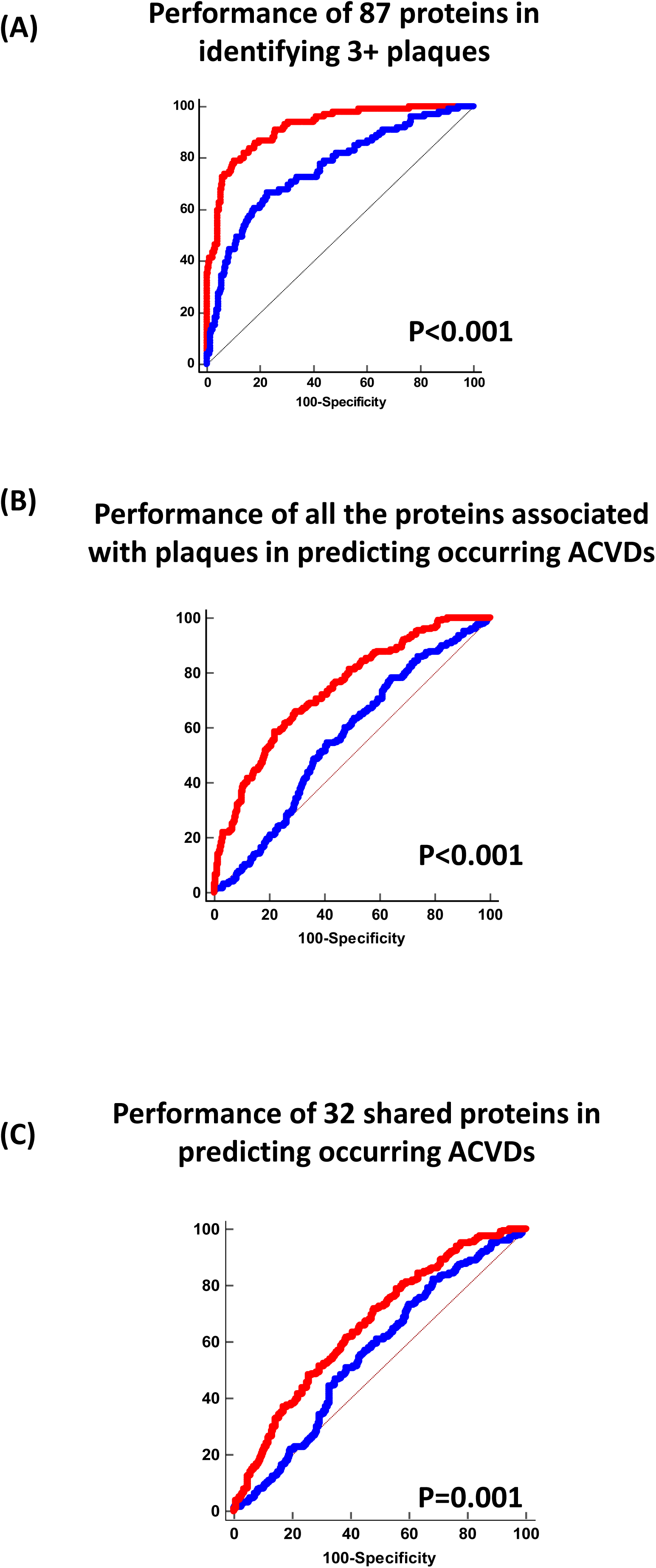
The plasma proteomics outperforms the ACVD risk factors in identifying subjects with 3+ plaques and in predicting occurring atherosclerotic events. Comparison of the accuracy (Receiver Operating Curve “ROC”) of the model to discriminate: (A) subjects with 3+ plaques from the controls, when including the ACVD risk factors together with the 87 proteins (including the 32 shared proteins) differently expressed in subjects with 3+ plaques (red curve) versus the accuracy of the model that included the ACVD risk factors only (blue curve); (B) subjects with incident ACVD, when including the ACVD risk factors and the 87 proteins associated with carotid plaques (including the 32 shared proteins) (red curve), versus the accuracy of the model that included the ACVD risk factors only (blue curve); (C) subjects with incident ACVD, when including the ACVD risk factors and the 32 shared proteins only (red curve), versus the accuracy of the model that included the ACVD risk factors only (blue curve); P values are derived from Hanley-McNeil test.

These findings support the concept that plasma proteomics independently predict the presence of a higher number of carotid plaques and improves our capability to identify apparently healthy subjects with diffuse carotid atherosclerosis (three or more plaques).

### Plasma proteins associated with number of carotid plaques also predict occurring ACVDs

Finally, to study whether the proteins that identify subjects with carotid plaques, and that recapitulate biologically relevant pathways in plasma, could also improve the performance of the commonly used risk factors to predict occurring ACVDs, we followed the population over 22 (21-23) years.

198 events occurred over time (29.81% in the population). Overall, ACVD classical risk factors modestly predicted the occurrence of the events (AUC= 0.559 (0.521-0.598)) and were not prevalent among subjects with an incurring event. By contrast, 87 proteins that were associated with the presence of 3+ plaques (including also the 32 shared proteins) markedly improved the ROC to predict the event vs the ROC of the model with the same classical risk factors only (AUC=0.739 (0.704-0.773), p<0.001; **Figure 4B**)).

Of note, the AUC of the ROC considering the 32 shared proteins only was also significantly higher vs the ROC considering the ACVD risk factors only (p=0.001), although its performance was attenuated (AUC= 0.660 (0.623-0.696); **Figure 4C**).

## Discussion

In this study we searched for proteins, out of a set of 368 measured in plasma by PEA technology, whose expressions can recapitulate biologically relevant pathways and can also improve the accuracy to identify apparently healthy subjects presenting with carotid plaques and to predict occurring ACVDs.

We detected proteins whose plasmatic expression significantly discriminates subjects who, although being apparently healthy according to the analysis of the classical ACVD risk factors, present with carotid atherosclerosis versus subjects who do not display atherosclerosis and that, in our study, were considered as “controls”. Moreover, we found that the significance of this association increased as a function of the number of carotid plaques analyzed by ultrasound, and we also provide a specific set of proteins that were associated with diffuse atherosclerosis (3+ plaques). Indeed, we identified 87 proteins, which associated with the presence of three or more plaques, presenting either a reduced (5) or increased (82) plasmatic expression compared to controls. By contrast, plasma proteomics was not able to significantly discriminate neither the subjects presenting with 2 or with just one carotid plaque. The finding that proteomics did not discriminate the subjects with lower number of plaques could perhaps be due to the smaller number of subjects in these groups. Anyhow, we cannot rule out the effect of the clinical conditions that mostly contribute to hemorheological turbulence and endothelial damage, such as hypertension and smoking habit, increased waist circumference, dyslipidemia and reduced physical activity, which were prevalent in subjects with 3+ plaques but not in the subjects with 2 or with 1 plaque only versus controls and which were also associated with higher NPX values for the majority of proteins analyzed (**Supplemental Figure 2A-E**).

The AUC of the model combining the information of these 87 proteins with that of the classical ACVD risk factors to discriminate subjects with 3+ plaques compared to controls was markedly elevated versus the AUC of the model considering the risk factors alone. Also, this significant improvement held true between a subset of the two groups, matched for prevalence of hypertension, active smoking habit, dyslipidemia, frequency of physical activity and similar waist circumference (AUC= 0.840 (0.771-0.895) of the model with proteins and ACVD risk factors vs AUC= 0.684 (0.602-0.757) with ACVD risk factors alone, p<0.001; not shown). Therefore, this observation further supports that plasma proteomics can improve our capability to classify, with higher accuracy, individuals among the apparently healthy population towards higher degrees of ACVD risk. Anyhow, we acknowledge that larger and independent studies are needed to validate this hypothesis.

We also reported comparable levels of CRP, a commonly considered marker of low grade inflammation in clinics, in subjects with 3+ plaques versus controls. We do not rule out the relevance of CRP, which is generally elevated in subjects at higher ACVD risk and co-morbid patients, but we are more likely to support the possibility that pre-clinical inflammation in “apparently healthy” individuals could be recaptured more significantly by alternative sets of proteins in plasma.

Furthermore, by combining a high-sensitive gradient boosting machine learning strategy together with the analysis of pathways, we also provided a biological association for the large number of proteins that were predicted to be linked to the number of carotid plaques. For instance, Growth Differentiation Factor 15 (GDF-15), whose NPX was increased in subjects with 3+ plaques versus controls, ranged first out of the 87 proteins in the random forest classifier (**Supplemental Table 7**), confirming independent epidemiological studies that previously associated its increased plasma levels to ACVD risk ^8,30^, to carotid atherosclerosis, increased carotid IMT ^7,9,10^ and high-risk coronary plaques defined by computed tomography angiography^31^. However, our additional GO analysis found that alternative set of proteins, including the spectrum of β-chemokines and other interleukins/cytokines, but not GDF-15, significantly clustered into biological pathways involved in the development of atherosclerotic process (**Supplemental Table 5** and **Supplemental table 6C**). From a translational point of view, these findings outline further mechanisms, involving immune cells chemotaxis in atherosclerotic lesions^32^ and lymphocytes activation in the atheroma^33^, taking place yet during the subclinical stages of atherosclerosis.

Finally, the robustness of our predictions is also supported by our follow-up analysis. Indeed, the 87 proteins (including the 32 shared proteins), identifying subjects with higher number of plaques, significantly predicted incident ACVDs over a large twenty-years follow-up. From a clinical standpoint, the finding that proteomics also markedly outperformed the accuracy of ACVD risk factors, further supports the possibility to overcome, by identifying individual proteomic fingerprints, the clinical algorithms which often underestimate the residual risk in apparently healthy subjects, owing a ten-year perspective for most of them.

Our study has some limitations, and some technical aspects of our work should be discussed.

First, our machine learning model, although previously demonstrated to provide superior accuracy compared to common statistical tools ^25^, was trained and applied in internal sets of our population and, consequently, our findings need to be validated in independent cohorts.

Second, the methodological assessment of our proteomics analysis, although providing an optimal sensitivity to target low proteins abundancy in plasma, otherwise not reachable by common techniques of molecular biology^34^, relies on PCR-based amplification reactions with antibodies-linked oligonucleotides. A quantitative validation of our results will add further strength; However, for some of the proteins we have identified, a validation by immunoassays has been provided^34^. All in all, compared existing literature using this strategy ^9,10,16^, we have provided information from one of the largest panels so far reported.

In conclusion, we found plasma proteins that, informing on pathways that are biologically relevant for the development of atherosclerosis, can identify apparently healthy subjects with higher number of plaques and that outperform the classical ACVD risk factors to predict occurring atherosclerotic events.

## Data Availability

The work includes data analyzed from an array of proteomics measured in plasma. All raw data can be provided upon reasonable written request to ALC.

## Acknowledgments

The work of A.B and A.L.C. is supported by a grant Ricerca Corrente from the Ministry of Health to IRCCS Multimedica; PRIN 2017H5F943; ERANET ER-2017-2364981 and a grant A.L.C. RF-2019-12370896

---

**Supplemental Figure 1.**
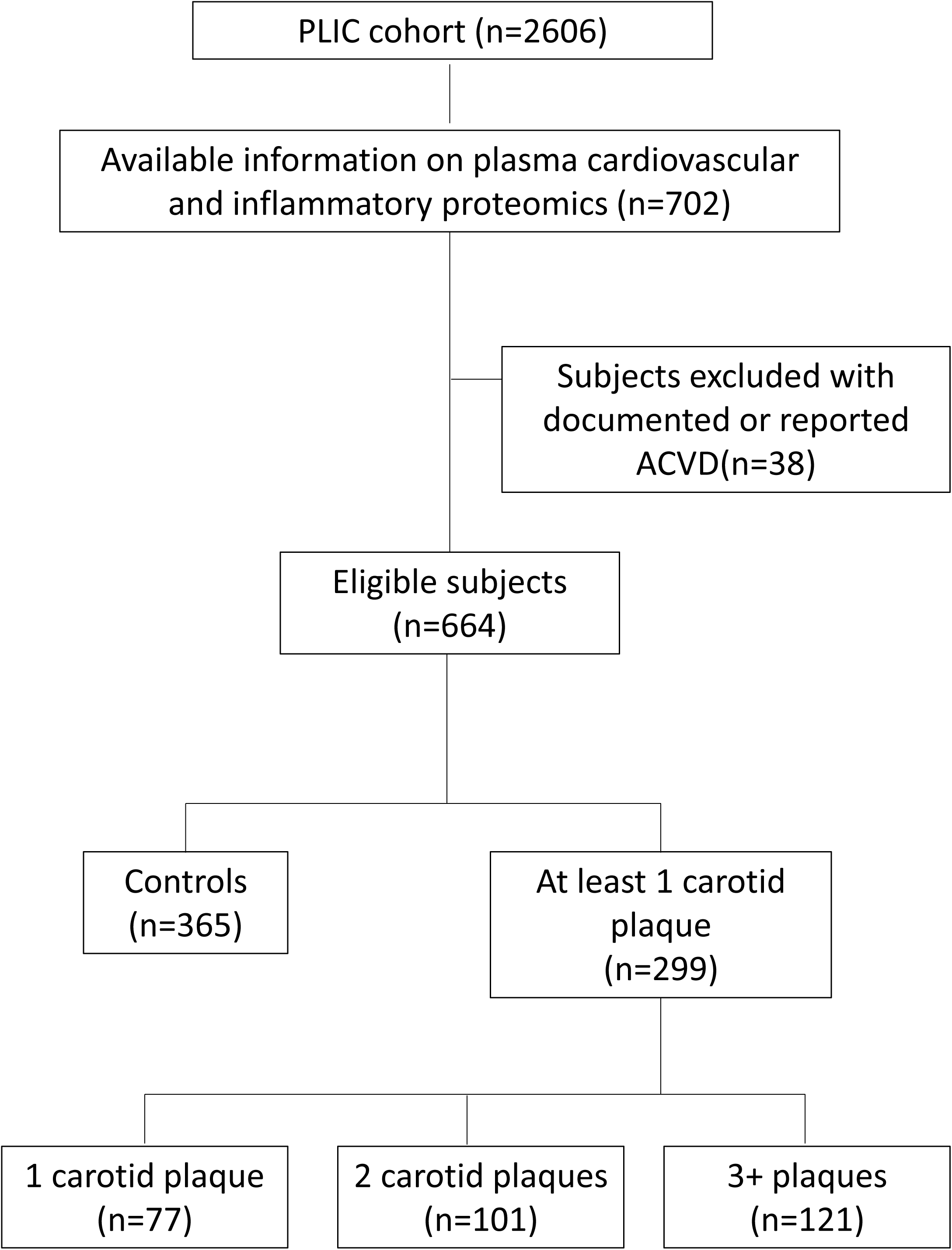
Study design.

**Supplemental Figure 2.**
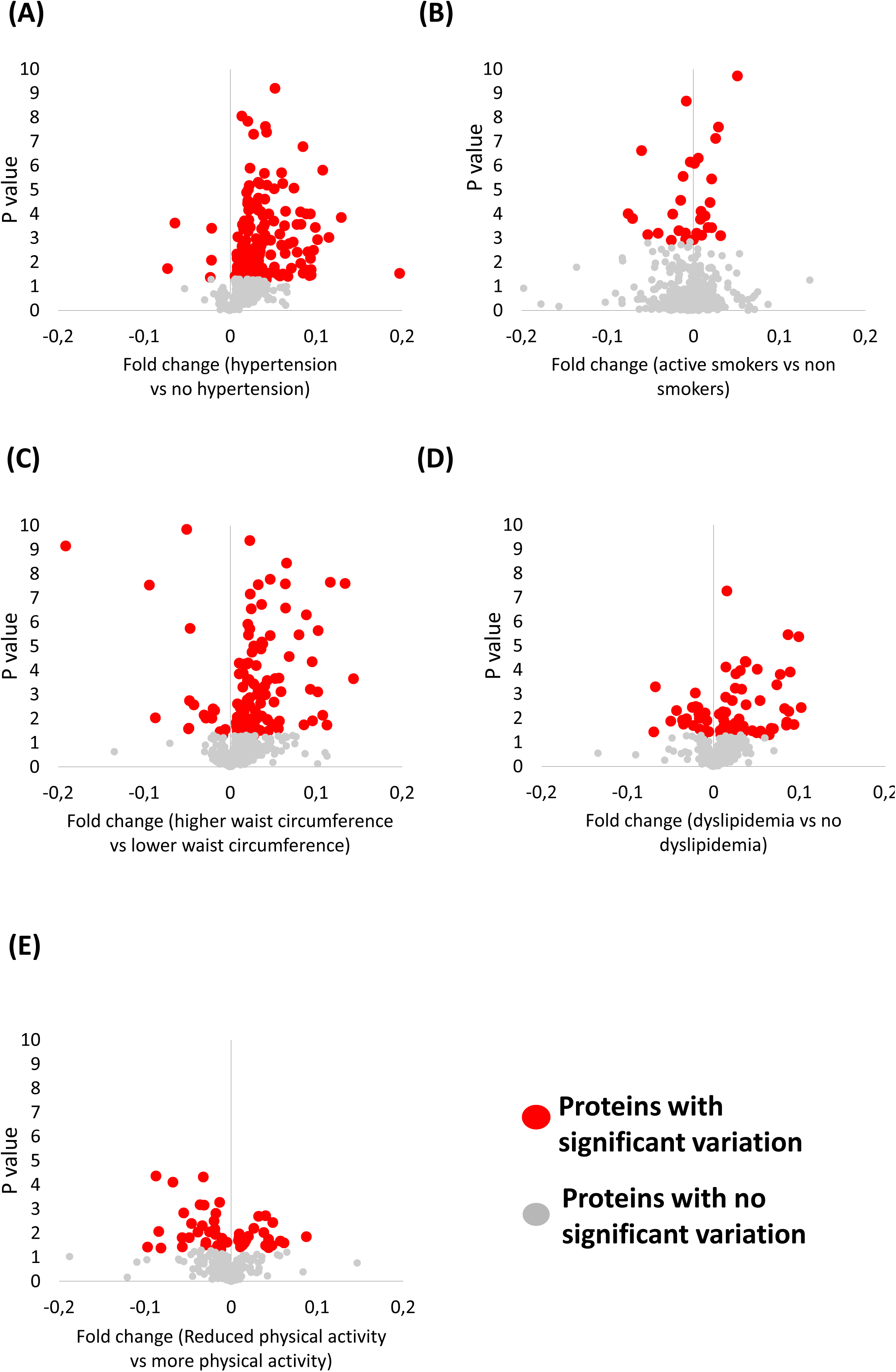
The plasmatic expression of several proteins associated with the ACVD risk factors. Volcano plots showing the variation of the 368 plasma proteins in **(A)** subjects with hypertension compared to subjects without hypertension, in **(B)** active smokers versus non-active smokers, **(C)** in subjects with higher waist circumference versus subjects with lower waist circumference, **(D)** in subjects with dyslipidemia and **(E)** in subjects who declare to practice less physical activity versus subjects who declared to practice more physical activity. All the variations are referred to controls and are calculated as fold of changes (x axis). In the y axis, the p value is reported indicating the significance of the variation.

